# “SARS-CoV-2 antibody seroprevalence and stability in a tertiary care hospital-setting”

**DOI:** 10.1101/2020.09.02.20186486

**Authors:** Samreen Siddiqui, Salwa Naushin, Shalini Pradhan, Archa Misra, Akansha Tyagi, Menka Loomba, Swati Waghdhare, Rajesh Pandey, Shantanu Sengupta, Sujeet Jha

**Affiliations:** Institute of Endocrinology, Diabetes, and Metabolism, Max Healthcare, Max Super Speciality Hospital, Saket,New Delhi, India; CSIR-Institute of Genomics and Integrative Biology, New Delhi-110025, India; Academy of Scientific and Innovative Research (AcSIR), Ghaziabad-201002, India

**Keywords:** SARS-CoV-2, seroprevalence, Healthcare workers (HCW), Asymptomatic, Antibody stability

## Abstract

**Background:** SARS-CoV-2 infection has caused 64,469 deaths in India, with 7, 81, 975 active cases till 30^th^ August 2020, lifting it to 3^rd^ rank globally. To estimate the burden of the disease with time it is important to undertake a longitudinal seroprevalence study which will also help to understand the stability of anti SARS-CoV-2 antibodies. Various studies have been conducted worldwide to assess the antibody stability. However, there is very limited data available from India. Healthcare workers (HCW) are the frontline workforce and more exposed to the COVID-19 infection (SARS-CoV-2) compared to the community. This study was conceptualized with an aim to estimate the seroprevalence in hospital and general population and determine the stability of anti SARS-CoV-2 antibodies in HCW.

**Methods:** Staff of a tertiary care hospital in Delhi and individuals visiting that hospital were recruited between April to August 2020. Venous blood sample, demographic, clinical, COVID-19 symptoms, and RT-PCR data was collected from all participants. Serological testing was performed using the electro-chemiluminescence based assay developed by Roche Diagnostics, in Cobas Elecsys 411. Seropositive participants were followed- upto 83 days to check for the presence of antibodies.

**Results:** A total of 780 participants were included in this study, which comprised 448 HCW and 332 individuals from the general population. Among the HCW, seroprevalence rates increased from 2.3% in April to 50.6% in July. The cumulative prevalence was 16.5% in HCW and 23.5% (78/332) in the general population with a large number of asymptomatic individuals. Out of 74 seropositive HCWs, 51 were followed-up for the duration of this study. We observed that in all seropositive cases the antibodies were sustained even up to 83 days.

**Conclusion:** **The** cumulative prevalence of seropositivity was lower in HCWs than the general population. There were a large number of asymptomatic cases and the antibodies developed persisted through the duration of the study. More such longitudinal serology studies are needed to better understand the antibody response kinetics.

## Introduction

In late December 2019, a group of novel coronavirus related pneumonia was reported from Wuhan, China^1^. On February 11, 2020 it was officially named by the World Health Organization (WHO) as coronavirus disease 2019 (COVID-19) caused by novel Severe Acute Respiratory Syndrome Coronavirus 2 (SARS-CoV-2)^2^. It has spread rapidly to more than 200 countries afflicting and challenging the health, economy and social well-being of millions of people. This widespread contagious disease was declared a world pandemic by WHO on 12th March 2020. The COVID-19 pandemic reached India in early 2020 with the first confirmed case in Kerala, while the first case in Delhi was reported on 3rd March 2020. With the rapid increase in the number of confirmed COVID-19 cases, India remains one of the severely affected countries with on-going pandemic. The magnitude of community spread has made Delhi a national epicentre with 1,69,412 cases and cumulative deaths of 4,389 till 29th August 2020^3^. With spread of infection through contact /aerosol exposure to the virus, it has been challenging to minimize community spread. Making the situation even worse is the spread of the virus by asymptomatic carriers, many of them unaware of being viral carriers. The situation is even more challenging for the Healthcare workers (HCWs) who have a greater chance of being infected given their occupational exposure^4^. For formulating public health policies and modifying the national response to COVID-19 pandemic, it is therefore crucial to recognize the risk of SARS-CoV-2 infection among the frontline medical staff and hospital system. This may provide a snapshot of current community spread of the virus.

Most of the individuals exposed to the virus develop antibodies within two to three weeks of exposure^5,6^. RT-PCR based molecular testing detects viral load during acute phase of infection which can control spread whereas serological tests could identify antibodies after acute infection and spot those cases that were missed by RT-PCR. However, it is important to assess the stability of the antibodies to estimate for how long an individual might be protected from re-infection. Several studies have been conducted to look at the seroprevalence of antibodies against SARSCoV-2 in different parts of the world^7-12^. They have reported the presence of antibodies among the asymptomatic individuals along with confirmed COVID-19 cases. Chen Y et al had reported 17.14% seropositivity among 105 healthcare workers in China during the epidemic peak^13^. Previously during many viral outbreaks, serological assessment in the community has proven to be useful in understanding the spread of the disease along with chances of development of herd immunity and previous exposure to virus^14-16^.

We conducted a prospective longitudinal observational study to estimate the prevalence of anti SARS-CoV-2 antibodies among workers of a private hospital in Delhi with different levels of exposure to COVID-19 cases. We also evaluated the seroprevalence among individuals from the general population who got tested there, to examine the community spread of COVID-19. Another key objective of this study was to figure out the stability of anti SARS-CoV-2 antibodies among the seropositive cases.

## Methods

This is a prospective observational study conducted in the last five months (April to August 2020). Ethics approval was taken from the Institutional Ethics Committee of Max Super Speciality Hospital, Saket, New Delhi (REF NO.:RS/MSSH/DDF/SKT-2/IEC/ENDO/20–12). Employees of Max hospital were approached by an email for participation in this study. Individuals from the general population visiting the hospital for COVID-19 testing were also recruited in the study. Written informed consent was obtained from all participants. Data collection form was filled, which consisted of demographic details, general health information, any possible symptoms related to COVID-19 and exposure to confirmed or suspected COVID-19 cases. In addition to symptoms related to COVID-19, implementation of hygiene and protection measures were also included in the questionnaire. Each participant was assigned a unique identification code along with the sample IDs. The serological testing was performed at the CSIR-Institute of Genomics and Integrative Biology (CSIR-IGIB) laboratory. A flow-chart of the study is presented in figure 1.

**Figure 1:**
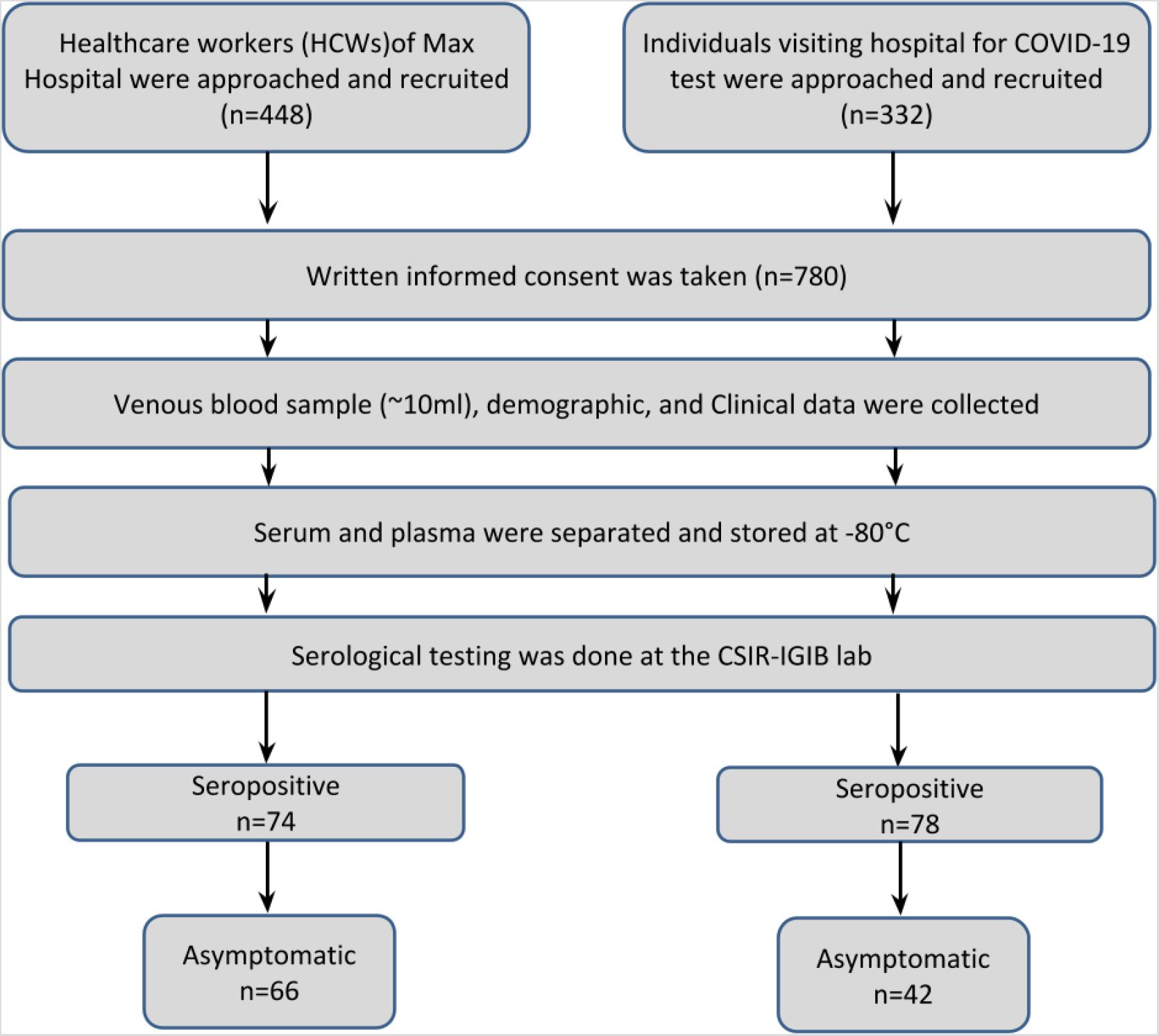
Workflow of the study.

### Blood sample collection

Institutional protocols recommending social distancing and screening of registered participants through thermal sensors at the time of sample collection were followed. Venous blood sample (∼10ml) was collected in 6ml EDTA and 5ml Serum Separating Tube (SST) from each participant. Serum and plasma were separated from them by centrifugation at 3000rpm for 15 minutes and stored at –80°C.

### COVID-19 serological testing

Plasma samples were used to run immunoassay tests for in vitro qualitative detection of antibodies against SARS-CoV-2, using electro-chemiluminescence based assay developed by Roche Diagnostics, in Cobas Elecsys 411, according to manufacturer’s protocol (Catalogue no. 92030958190), Roche anti SARS-Cov-2 kit, Roche Diagnostic^17^.

Participants who were found seropositive were contacted for follow-up and their blood samples were collected for checking the stability of the anti SARS-CoV-2 antibodies. Days of follow-up were calculated from the date they were first tested positive for the antibody.

### Statistical analyses

Seroprevalence in our sample sets was determined by the fractions of samples which tested positive with the commercially available immunoassay kit. We also stratified our analysis on the basis of age, gender, type of occupation and any proximity of the participant with COVID-19 positive case. We have also tried to anticipate the relation between the number of COVID-19 antibodies among the seropositive cases. symptoms experienced and seroconversion rate. The proportion of individuals who developed antibodies after being positively tested by RT-PCR was also looked upon. Continuous data like age is presented as mean±SD and categorical data such as gender, occupation, symptomatic/asymptomatic status, etc. are presented as percentages and number.

## Results

A total of 780 samples were included for the serological testing which included hospital workers and individuals visiting hospital during the pandemic. In total, 448 staff [physicians (n = 59), nurses (n = 70), administrative (n = 15), front office (n = 12), catering (n = 17), housekeeping (n = 46), security (n = 9), laboratory (n = 45), pharmacy (n = 8), general duty assistant (n = 28), engineering (n = 21), homecare (n = 5), research (n = 19), and others (n = 94)] from different hospital units agreed to participate in the study. There were 65.8% males and 34.1% females among the hospital staff. 332 individuals from the general population (77.1% males and 22.9% females) also participated in the study. The mean (SD) age of the healthcare workers was 32.5±9.8 years and that of the general population was 39.6±13.1 years. Demographic details of the two population subsets are presented in table 1 and 2, respectively.

**Table 1.**
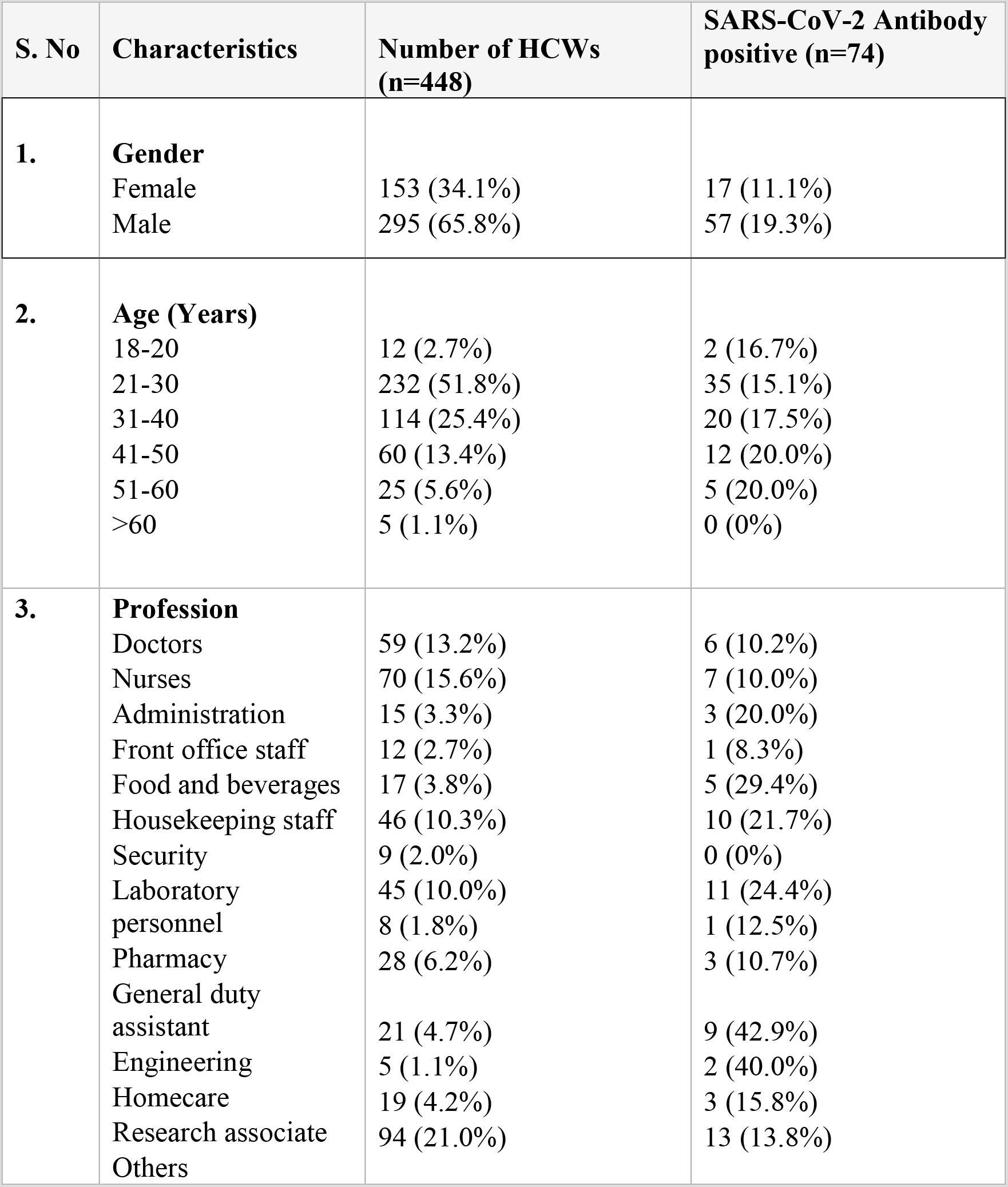
General characteristics and SARS-CoV-2 antibody positivity of the healthcare workers.

**Table 2.** General characteristics and SARS-CoV-2 antibody positivity of individuals from the general population.

| S.No | Characteristics | Number of participants from general population (n=332) | SARS-CoV-2 Antibody positive (n=78) |
| --- | --- | --- | --- |
| 1. | **Gender** |  |  |
|  | Female | 76 (22.9%) | 19 (25.0%) |
|  | Male | 256 (77.1%) | 59 (23.0%) |
| 2. | **Age (Years)** |  |  |
|  | 18-20 | 7 (2.1%) | 1 (14.3%) |
|  | 21-30 | 79 (23.8%) | 11 (13.9%) |
|  | 31-40 | 90 (27.1%) | 22 (24.4%) |
|  | 41-50 | 79 (23.8%) | 22 (27.8%) |
|  | 51-60 | 49 (14.8%) | 14 (28.6%) |
|  | >60 | 28 (8.4%) | 8 (28.6%) |
| 3. | **Profession** |  |  |
|  | Government employee | 35 (10.5%) | 7 (20.0%) |
|  | Private Job | 47 (14.2%) | 11 (23.4%) |
|  | Business | 23 (6.9%) | 6 (26.1%) |
|  | Manual-skilled | 16 (4.8%) | 1 (6.2%) |
|  | Food services | 8 (2.4%) | 3 (37.5%) |
|  | Student | 14 (4.2%) | 3 (21.4%) |
|  | Delhi Police | 27 (8.1%) | 2 (7.4%) |
|  | Driver | 9 (2.7%) | 1 (11.1%) |
|  | Homemaker | 37 (11.1%) | 7 (18.9%) |
|  | Teaching | 8 (2.4%) | 2 (25.0%) |
|  | Vegetable seller | 4 (1.2%) | 1 (25.0%) |
|  | Retired | 10 (3.0%) | 2 (20.0%) |
|  | Unknown | 57 (17.2%) | 27 (47.4%) |
|  | Others | 37 (11.1%) | 5 (13.5%) |

**Table 3.** COVID-19-related symptoms of the healthcare workers.

| S.No |  | Number of HCWs<br>(n=448) | SARS-CoV-2<br>Antibody positive<br>(n=74) |
| --- | --- | --- | --- |
| 1. | **Presence of COVID-19 related symptoms**<br>Asymptomatic<br>Symptomatic | 374 (83.5%)<br>74 (16.5%) | 66 (17.5%)<br>8 (10.7%) |
| 2. | **Common symptoms**<br>Fever<br>Cough<br>Sore throat<br>Breathlessness<br>Myalgia<br>Others | 43 (9.6%)<br>38 (8.5%)<br>47 (10.5%)<br>13 (2.9%)<br>6 (1.3%)<br>62 (13.8%) | 4 (9.3%)<br>8 (21.0%)<br>4 (8.5%)<br>1 (7.7%)<br>1 (16.7%)<br>10 (16.1%) |
| 3. | **Physical proximity with COVID-19 positive**<br>Yes<br>No<br>Maybe | 184 (41.1%)<br>95 (21.2%)<br>169 (37.7%) | 43 (23.4%)<br>7 (7.4%)<br>24 (14.2%) |
| 4. | **Contact with symptomatic/suspected person**<br>Yes<br>No<br>Maybe | 18 (4.1%)<br>188 (42.0%)<br>242 (54.0%) | 7 (38.9%)<br>21 (11.2%)<br>46 (19.0%) |

**Table 4.**
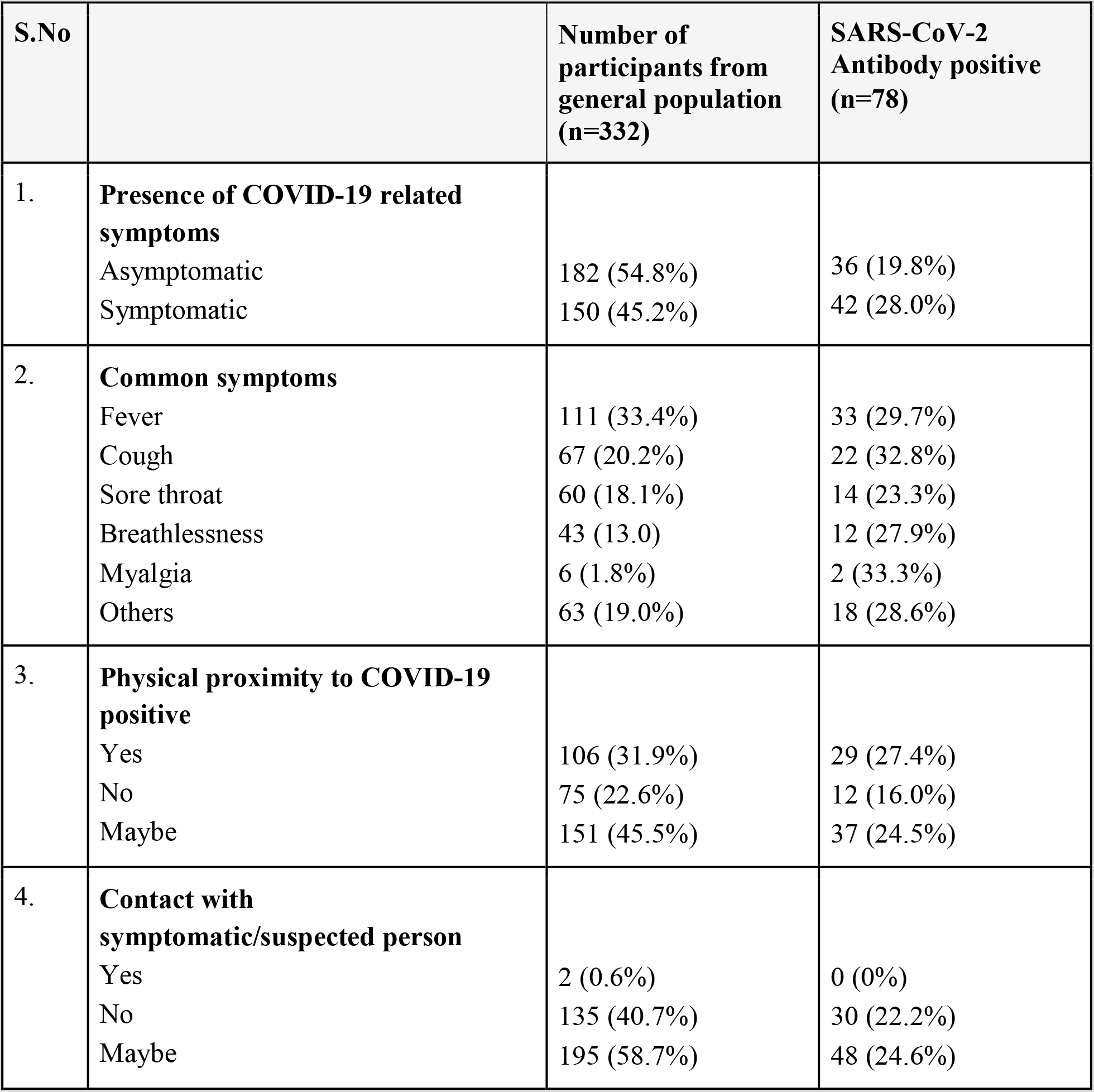
COVID-19-related symptoms of the general population.

| S.No |  | Number of participants from general population (n=332) | SARS-CoV-2 Antibody positive (n=78) |
| --- | --- | --- | --- |
| 1. | **Presence of COVID-19 related symptoms**<br>Asymptomatic<br>Symptomatic | 182 (54.8%)<br>150 (45.2%) | 36 (19.8%)<br>42 (28.0%) |
| 2. | **Common symptoms**<br>Fever<br>Cough<br>Sore throat<br>Breathlessness<br>Myalgia<br>Others | 111 (33.4%)<br>67 (20.2%)<br>60 (18.1%)<br>43 (13.0%)<br>6 (1.8%)<br>63 (19.0%) | 33 (29.7%)<br>22 (32.8%)<br>14 (23.3%)<br>12 (27.9%)<br>2 (33.3%)<br>18 (28.6%) |
| 3. | **Physical proximity to COVID-19 positive**<br>Yes<br>No<br>Maybe | 106 (31.9%)<br>75 (22.6%)<br>151 (45.5%) | 29 (27.4%)<br>12 (16.0%)<br>37 (24.5%) |
| 4. | **Contact with symptomatic/suspected person**<br>Yes<br>No<br>Maybe | 2 (0.6%)<br>135 (40.7%)<br>195 (58.7%) | 0 (0%)<br>30 (22.2%)<br>48 (24.6%) |

Among the HCWs, the seroprevalence increased from April (2.3%) to July (50.6%) as expected (Figure 2). The cumulative seroprevalence observed in our study is 16.5% (74/448). The prevalence observed in the general population is 23.5% (78/332). The prevalence of seropositivity was higher in males in case of hospital workers (19.3% vs 11.1%) whereas no such difference was found in the other group (23.0% in males vs 25.0% in females). Out of those tested positive for the antibody in the study, 67.1% (102/152) were asymptomatic while 32.9% (50/152) had COVID-19 related symptoms like fever (n = 37), sore throat (n = 18), cough (n = 30), breathlessness (n = 13), myalgia (n = 3), and other mild symptoms (n = 28) including headache, abdominal pain. 48.0% (49/102) of the asymptomatic participants, who were tested seropositive, were not reported to have been exposed to any confirmed or suspected case of COVID-19.

**Figure 2.**
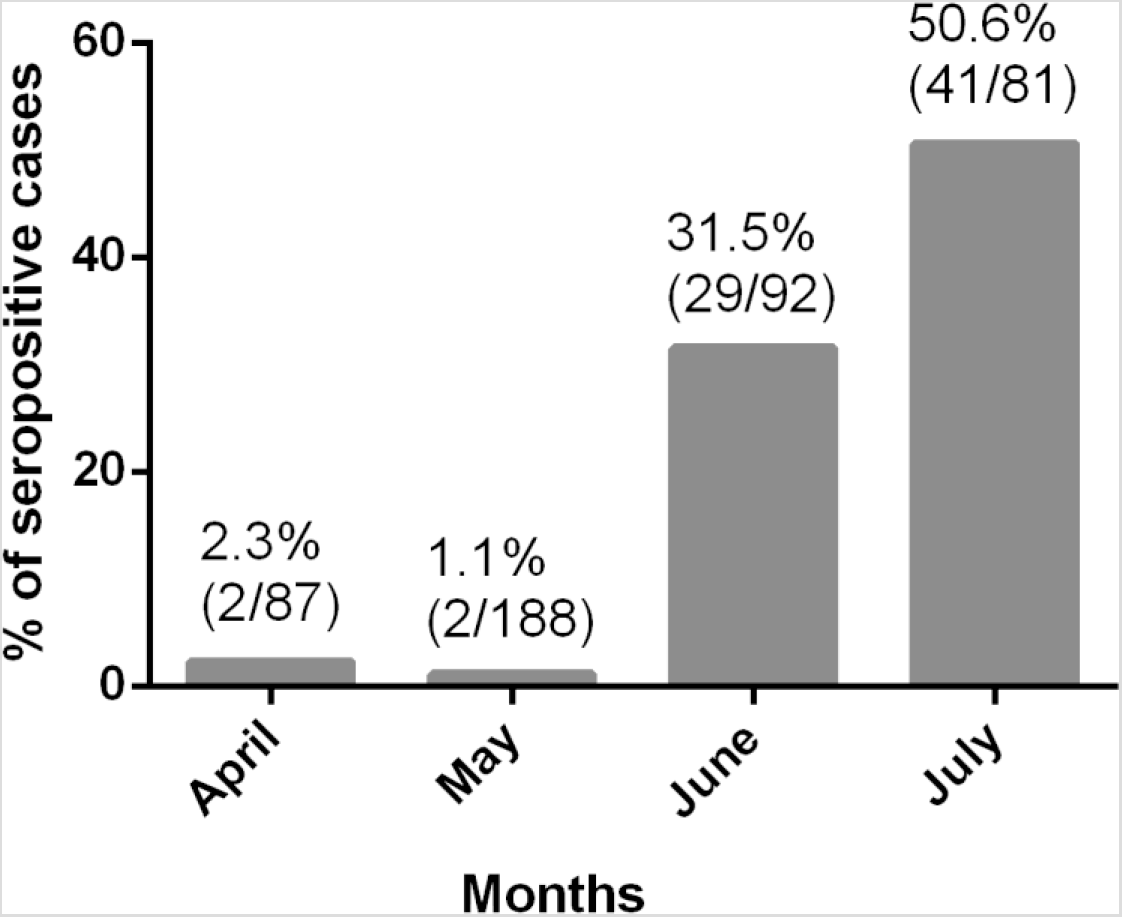
Month-wise seroprevalence in healthcare workers.The percentage is shown in the top below which seropositive cases/total no. of participants are mentioned.

On comparing the two population subsets we observed that 89.2% (66/74) of the seropositive HCWs were asymptomatic compared to 46.1% (36/78) individuals of the general population. Interestingly, among the healthcare workers, doctors (6/59, 10.2%) and nurses (7/72, 9.7%) had lower seropositivity rates than the other staff, engineering (9/21, 42.9%), food and beverages (5/17, 29.4%), the laboratory staff (11/45, 24.4%), and the housekeeping staff (10/46, 21.7%). For the general population, seropositivity varied across different occupations (table 2).

### Stability of anti SARS-CoV-2 antibodies

Of the 74 HCWs who tested positive for the anti SARS-CoV-2 antibody, 51 participants were followed-up for various time intervals, and presence of antibodies was assessed at intervals of 7–15 days using the same assay protocol (table 5). It was observed that antibodies against SARSCoV-2 were sustained in these participants for more than 60 days with the longest persistence of 83 days in one of the participants (figure 3a-3d). Among these, 19 participants had sustained antibody levels at least until 40 days after the first detection.

**Table 5.** Follow-up data of antibodies in seropositive participants.

| S.No. | Samples | Day 1 | 7-14 days | 15-30 days | 31-45 days | 46-60 days | 61-80 days | 81-95 days |
| --- | --- | --- | --- | --- | --- | --- | --- | --- |
| 1 | ID0001 | 1.3 |  |  | 19.6 |  |  | 16.7 |
| 2 | ID0002 | 2.3 | 1.7 |  | 2 |  | 1.9 |  |
| 3 | ID0003 | 68.7 |  | 67.5 | 61.3 |  | 56 |  |
| 4 | ID0004 | 7 | 12.7 |  |  |  | 124.5 |  |
| 5 | ID0005 | 6.1 |  |  | 24.3 |  |  |  |
| 6 | ID0006 | 19.8 |  |  |  | 58.7 |  |  |
| 7 | ID0007 | 57.5 |  |  | 123.3 |  |  |  |
| 8 | ID0008 | 4.6 |  | 72.5 |  |  |  |  |
| 9 | ID0009 | 2 |  | 13.6 |  |  |  |  |
| 10 | ID0010 | 1.8 |  | 36.5 |  |  |  |  |
| 11 | ID0011 | 0.9 | 2.2 | 2.1 |  | 3 |  |  |
| 12 | ID0012 | 4.1 |  |  |  | 27.6 |  |  |
| 13 | ID0013 | 2.4 | 7.9 |  | 22.4 |  |  |  |
| 14 | ID0014 | 5.5 | 15.8 |  |  |  |  |  |
| 15 | ID0015 | 33.8 | 80.7 |  | 95.3 |  |  |  |
| 16 | ID0016 | 7.3 |  | 17.3 |  | 59.9 |  |  |
| 17 | ID0017 | 6.7 |  |  | 3.9 |  |  |  |
| 18 | ID0018 | 2.4 |  | 20 |  | 50.8 |  |  |
| 19 | ID0019 | 3.7 |  | 12.9 |  |  |  |  |
| 20 | ID0020 | 3.8 |  | 65.8 |  |  |  |  |
| 21 | ID0021 | 1.6 |  |  |  | 18.9 |  |  |
| 22 | ID0022 | 6.3 |  | 6.8 |  |  |  |  |
| 23 | ID0023 | 20.4 | 27.5 |  |  |  |  |  |
| 24 | ID0024 | 1.1 | 1.6 |  | 26.4 |  |  |  |
| 25 | ID0025 | 2.5 |  | 65.5 |  |  |  |  |
| 26 | ID0026 | 11 |  |  |  | 43.9 |  |  |
| 27 | ID0027 | 8 | 4.7 |  |  |  |  |  |
| 28 | ID0028 | 5.7 |  | 21.9 |  |  |  |  |
| 29 | ID0029 | 27.2 |  |  |  |  |  |  |
| 30 | ID0030 | 3.4 |  | 24.9 |  |  |  |  |
| 31 | ID0031 | 16.6 |  | 29.4 |  |  |  |  |
| 32 | ID0032 | 8.8 |  |  |  | 77.6 |  |  |
| 33 | ID0033 | 1 |  |  |  | 78 |  |  |
| 34 | ID0034 | 7.2 |  | 11.5 | 12.7 |  |  |  |
| 35 | ID0035 | 1 |  |  |  | 57.3 |  |  |
| 36 | ID0036 | 1.6 |  |  |  | 34.1 |  |  |
| 37 | ID0037 | 1 | 2.9 |  | 9.4 |  |  |  |
| 38 | ID0038 | 31.3 |  |  | 40.7 |  |  |  |
| 39 | ID0039 | 12.3 |  |  | 93.2 |  |  |  |
| 40 | ID0040 | 2.1 |  | 6.7 |  |  |  |  |
| 41 | ID0041 | 16.3 |  | 45.5 |  |  |  |  |
| 42 | ID0042 | 12.7 |  | 109.7 |  |  |  |  |
| 43 | ID0043 | 105.7 |  |  |  | 104 |  |  |
| 44 | ID0044 | 3.3 |  |  | 15.8 |  |  |  |
| 45 | ID0045 | 77 |  |  | 122.3 |  |  |  |
| 46 | ID0046 | 5.92 |  |  | 12 |  |  |  |
| 47 | ID0047 | 67.1 |  |  | 119.6 |  |  |  |
| 48 | ID0048 | 97.1 |  |  | 115.2 |  |  |  |
| 49 | ID0049 | 1.3 |  | 2.4 |  |  |  |  |
| 50 | ID0050 | 3.4 |  | 7.1 | 22 |  |  |  |
| 51 | ID0051 | 4.6 |  | 32.1 |  |  |  |  |

**Figure 3:**
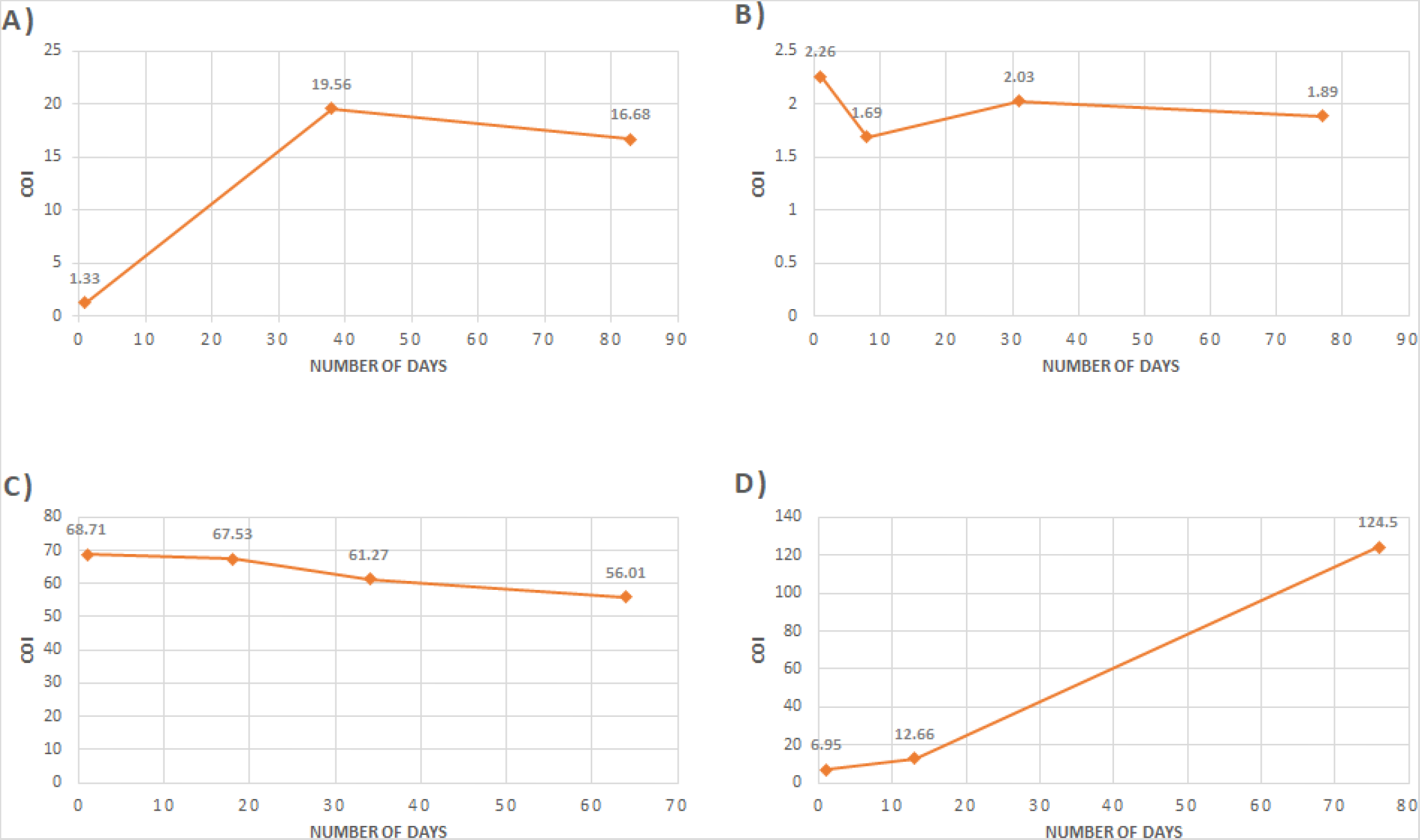
Follow-up data of participants more than 60 days. A) Participant 1, B) Participant 2, C) Participant 3 and D) Participant 4. COI represents Cut-off Index.

## Discussion

Healthcare workers are at the frontline in this pandemic and presumably more exposed to the virus than the community^4,8^. Assessing the antibody prevalence and its stability among seropositive individuals would help in developing public policies and estimating the risk of disease spread within a healthcare system. This kind of serology study could be useful to develop understanding regarding the efficacy of vaccines in clinical trials. Also, identifying individuals with high seroconversion rates can be beneficial for developing convalescent plasma therapy. The present study was conducted to estimate the seroprevalence of SARS-CoV-2 infection in Delhi and to observe how long the antibodies persist in the body. We included the HCWs of a private institute and individuals from the general population in this study. We analyzed plasma/serum samples for antibodies detection through electrochemiluminescence immunoassay using Roche kit, which has been already validated in our lab. In a separate study, we have found that the sensitivity and specificity of this kit was 94.5% and 99.4%, respectively (unpublished data).

We observed an increasing trend of seropositive cases amongst the hospital staff, over a period of four months, from April to July which was expected and is a reflection of the increased spread of the infection in these months. Interestingly, there were a large number of asymptomatic individuals who were found to be seropositive (17.6% in HCW, 19.8% in the general population). This is in agreement with what has been found in other parts of the world^10,18,19^. Studies in the U.K and Spain reported 10.6% and 23.1% seroprevalence among the HCW^7,8^. Recently, the National Center for Disease Control (NCDC) in collaboration with the Delhi government reported a 23.48% seropositive rate in the city through the study conducted between 27^th^ June-10^th^ July 2020^20^. One of the major limitations of this survey was that it was done over a very short period of time during the whole pandemic period. Our data on seropositivity amongst individuals visiting the hospital (23.5%) is in concordance with the Delhi sero survey. However, the seroprevalence was lower amongst healthcare workers (16.5%). Interestingly, even among the healthcare workers, doctors and nurses who are actually in close proximity to the COVID-19 cases had lower seropositivity. Although counterintuitive, it can be perceived that healthcare workers in general and doctors and nurses in particular strictly follow all the guidelines that help in protecting them against infection.

Several studies have been conducted globally to estimate the seroconversion rate among the HCW as well as the general population, including both symptomatic and asymptomatic individuals^7-12^. However, only a few studies have been done on antibody stability, including the population-based seroepidemiological study conducted in Spain which reported ∼90% seroprevalence after 14 days since the positive RT-PCR^18^. A multi-centre cohort study in the U.K tested HCWs and observed 47% seropositivity at more than 14 days after onset of symptoms^5^. Long et al, in China reported that the levels of neutralizing antibodies against SARSCoV-2, in those recovered from the disease, start decreasing after 2–3 months of infection^21^. Another such study noticed anti-SARS-CoV-2 IgM in one of the cases at 42 days after positive RT-PCR^22^. However, there is very limited data available on the antibody stability from India so far. We successfully followed-up a few of the seropositive participants and observed that antibodies against the infection last for 60–80 days, which is the maximum duration of follow up that could be done. These individuals will continue to be followed up. This might indicate the individual specific variation in the seroconversion rate of this virus infection. Questioning the seroprotection, a recent report has confirmed COVID-reinfection in a Hong Kong citizen, who was tested RT-PCR positive four-and-a-half months after being recovered from the disease^23^. Still much detail is not known about the immunogenic response due to this virus and needs further research to make definitive conclusions.

In a recently published brief report from Mumbai, India, conducted among the HCWs of three hospitals, highlighted that SARS-CoV-2 antibodies are not detected after 50 days, in RT-PCR positive individuals contrasting our observations^24^. This difference in the results might be due to differences in the population structure of Delhi and Mumbai and also probably due to the different strains of SARS-CoV-2 virus prevalent in the two cities^25^.

One of the major strengths of our study is that we benefitted from a longitudinal study over a period of 5 months with follow-up sampling from same individuals facilitating insights into the duration of seropositivity. This would be the first such preliminary report from India which provides evidence that the anti SARS-CoV-2 antibodies, once developed, could persist in the body for more than 60 days. Secondly, our findings suggest that antibodies can be developed in asymptomatic individuals without even being exposed to any confirmed or suspected case of COVID-19. This study has highlighted the importance of screening individuals irrespective of the presence or absence of COVID-19 related symptoms.

We recognize that there are a few limitations of this study. Firstly, the sample size was not large enough to generalize the study findings to a population. Additionally, the antibody stability was assessed for up to a maximum of 83 days. Observing the levels of antibodies for a longer duration could be helpful in getting more conclusive results. This might give insight about SARS-CoV-2 antibody response kinetics.

In conclusion, our study results confirm that anti SARS-CoV-2 antibodies could remain for more than 60 days in the body. This is a step forward towards better understanding of the infection recovery and re-infection pattern. There is a need for larger follow-up studies to further assess how long the antibodies remain stabilized in the body. Seroprevalence in our study expectedly increased from April to July with a cumulative rate of 16.5% among the HCWs, and 23.5% among the general population. We observed a higher rate of seroprevalence among the asymptomatic individuals. Larger population-based studies might be helpful in evaluating the immune response of the antibodies against re-infection.

## Data Availability

all data relavent to the manuscrpt are with us.

## Acknowledgements

Authors would like to acknowledge Dr. Mitali Mukerji, Chief Scientist, CSIR-IGIB for facilitating this collaborative research project. We would also like to acknowledge CSIR Project MLP 2003 (Combined Digital Surveillance and Effective COVID-19 Testing Frameworks and Tools) for funding this work. SN acknowledges CSIR for fellowship. RP acknowledges the funding from Fondation Botnar (CLP-0031) and CSIR (MLP-2005) towards the study.

## Author’s Contribution

SJ, SS*, SW conceptualized the study. SS* supervised the sample analysis, reviewed and edited the manuscript. SJ, SS#, SW supervised sample and data collection.SN analysed the samples and data. SN and SS# drafted the manuscript. SP analysed the samples and data.AM, AT, and ML collected samples and data. SW supervised sample and data collection. RP reviewed the manuscript.

